# DirEct Versus VIdeo LaryngosCopE (DEVICE): Protocol and statistical analysis plan for a randomized clinical trial in critically ill adults undergoing emergency tracheal intubation

**DOI:** 10.1101/2022.11.07.22282046

**Authors:** Matthew E. Prekker, Brian E. Driver, Stacy A. Trent, Daniel Resnick-Ault, Kevin P. Seitz, Derek W. Russell, Sheetal Gandotra, John P. Gaillard, Kevin W. Gibbs, Andrew J. Latimer, Micah R. Whitson, Shekhar A. Ghamande, Derek J. Vonderhaar, Jeremy P. Walco, Sydney J. Hansen, Ivor S. Douglas, Christopher R. Barnes, Vijay Krishnamoorthy, Jill J. Bastman, Bradley D. Lloyd, Sarah W. Robison, Jessica A. Palakshappa, Steven H. Mitchell, David B. Page, Heath D. White, Alyssa Espinera, Christopher G. Hughes, Aaron Joffe, J. Taylor Herbert, LTC Steven G. Schauer, Maj. Brit J. Long, Brant Imhoff, Li Wang, Jillian P. Rhoads, Kelsey N. Womack, David R. Janz, Wesley H. Self, Todd W. Rice, Adit A. Ginde, Jonathan D. Casey, Matthew W. Semler, the DEVICE investigators and the Pragmatic Critical Care Research Group

## Abstract

**Introduction:** Among critically ill patients undergoing orotracheal intubation in the emergency department (ED) or intensive care unit (ICU), failure to visualize the vocal cords and intubate the trachea on the first attempt is associated with an increased risk of complications. Two types of laryngoscopes are commonly available: direct laryngoscopes and video laryngoscopes. For critically ill adults undergoing emergency tracheal intubation, it remains uncertain whether use of a video laryngoscope increases the incidence of successful intubation on the first attempt compared with use of a direct laryngoscope.

**Methods and Analysis:** The <u>D</u>ir<u>E</u>ct Versus <u>VI</u>deo Laryngos<u>C</u>op<u>E</u> (DEVICE) trial is a prospective, multi-center, non-blinded, randomized trial being conducted in 6 EDs and 10 ICUs in the United States. The trial plans to enroll up to 2,000 critically ill adults undergoing orotracheal intubation with a laryngoscope. Eligible patients are randomized 1:1 to the use of a video laryngoscope or a direct laryngoscope for the first intubation attempt. The primary outcome is successful intubation on the first attempt. The secondary outcome is the incidence of severe complications between induction and 2 minutes after intubation, defined as the occurrence of one or more of the following: severe hypoxemia (lowest oxygen saturation < 80%); severe hypotension (systolic blood pressure < 65 mm Hg or new or increased vasopressor administration); cardiac arrest; or death. Enrollment began on March 16, 2022 and is expected to be completed in 2023.

**Ethics and Dissemination:** The trial protocol was approved with waiver of informed consent by the single institutional review board at Vanderbilt University Medical Center and the Human Research Protection Office of the Department of Defense. The results will be presented at scientific conferences and submitted for publication in a peer-reviewed journal.

**Trial Registration:** ClinicalTrials.gov registration (NCT05239195) on February 14, 2022, prior to the enrollment of the first patient.

**Strengths and Limitations of this Study:**

- This protocol describes in detail the design and methods for a large, pragmatic trial of laryngoscope type for the emergency tracheal intubation of critically ill adults.
- Conduct in the emergency departments and intensive care units of multiple centers among operators with diverse prior experience with tracheal intubation, as well as broad patient eligibility criteria, will increase the external validity of trial results.
- Patients, clinicians, and investigators are not blinded to the study group assignment after randomization.

## Introduction

Tracheal intubation is a common procedure in the emergency department (ED) and intensive care unit (ICU). Among critically ill patients undergoing tracheal intubation, failure to intubate the trachea on the first attempt is associated with increased risk of complications, including hypoxemia, hypotension, aspiration, and cardiac arrest.^1,2^

Emergency tracheal intubation is typically performed in three discrete steps. First, the patient is administered medications to facilitate optimal intubating conditions (rapid sequence induction). Second, a clinician inserts a laryngoscope into the patient’s mouth to visualize the vocal cords (laryngoscopy). Third, an endotracheal tube is inserted into the mouth, alongside the laryngoscope, and the tube is advanced past the vocal cords into the trachea (intubation).

The direct laryngoscope, the traditional instrument consisting of a battery-containing handle attached to a blade with a light source, has been used to visualize the vocal cords for tracheal intubation for over 100 years and remains the most commonly used device for the intubation of critically ill adults in the ED or ICU.^2–5^ The operator uses the direct laryngoscope to displace the tongue and elevate the epiglottis to facilitate intubation of the trachea under direct visualization. Obtaining an adequate view of the larynx with a direct laryngoscope can be challenging, especially for inexperienced operators. Once a view of the larynx is obtained, passage of the endotracheal tube follows the operator’s direct line-of-sight through the mouth to the vocal cords.

Over the last two decades, video laryngoscopes have provided an alternative to direct laryngoscopes for visualizing the vocal cords to facilitate tracheal intubation.^6,7^ A camera embedded near the tip of the video laryngoscope blade transmits an image of the vocal cords to a screen that the operator can view during the procedure.^8^ Because the camera is located near the tip of the laryngoscope blade, obtaining a view of the larynx may be easier with a video laryngoscope compared with a direct laryngoscope. However, because this view can be obtained without generating a direct line-of-sight through the mouth to the vocal cords, the process of passing an endotracheal tube may be more difficult when using a video laryngoscope. When considering both aspects of tracheal intubation, visualizing the vocal cords and passing the endotracheal tube, it remains uncertain whether use of a video laryngoscope increases the incidence of successful intubation on the first attempt.

Among elective tracheal intubations in the operating room, use of video laryngoscope probably increases the incidence of successful intubation on the first attempt and decreases complications compared to use of a direct laryngoscope, supported with moderate certainty in the existing anesthesiology literature.^9^ Extrapolating the results of randomized clinical trials conducted in the operating room to non-operating room settings is problematic because of factors related to the patient, the operator, and the environment.^10,11^ Because tracheal intubation of critically ill adults outside of the operating room is common, complications of intubation in the ED and ICU are common, and use of a video laryngoscope during intubation in the ED and ICU has increased significantly over time,^9,12^ understanding the effects of use of a video laryngoscope vs direct laryngoscope on successful intubation on the first attempt in these settings is a priority.

Previous trials randomizing patients to use of a video laryngoscope or a direct laryngoscope during emergency tracheal intubation in prehospital^13–18^, ED^19–25^, and ICU settings^26–32^ have been small and heterogeneous and have generally suggested that while a video laryngoscope improves the view of the larynx and reduces the incidence of esophageal intubation, it may not affect the incidence of successful intubation on the first attempt. Findings were similar in the largest such trial to date, a 371-patient, multicenter, randomized clinical trial in French medical ICUs in which use of video laryngoscope failed to improve successful intubation on the first attempt (68% vs. 70%; p = 0.60) and was associated with a greater incidence of severe peri-procedural complications in post-hoc analyses.^33^

The sample size of these prior trials did not provide sufficient statistical power to definitively rule out a clinically important effect of use of a video laryngoscope vs direct laryngoscope on successful intubation on the first laryngoscopy attempt or the incidence of complications. To compare the effectiveness of these two commonly used devices during this important emergency procedure, a large trial conducted across a wide variety of clinical settings, operator specialties, and levels of operator experience is required. Therefore, we designed the <u>D</u>ir<u>E</u>ct Versus <u>VI</u>deo Laryngos<u>C</u>op<u>E</u> (DEVICE) trial to test the hypothesis that, among critically ill adults undergoing emergency tracheal intubation in the ED or ICU, use of a video laryngoscope will increase the incidence of successful intubation on the first attempt compared with use of a direct laryngoscope.

## Methods and Analysis

This manuscript was written in accordance with Standard Protocol Items: Recommendations for Interventional Trials (SPIRIT) guidelines (Figure 1; supplementary file, section 1).^34^

**Figure 1.**
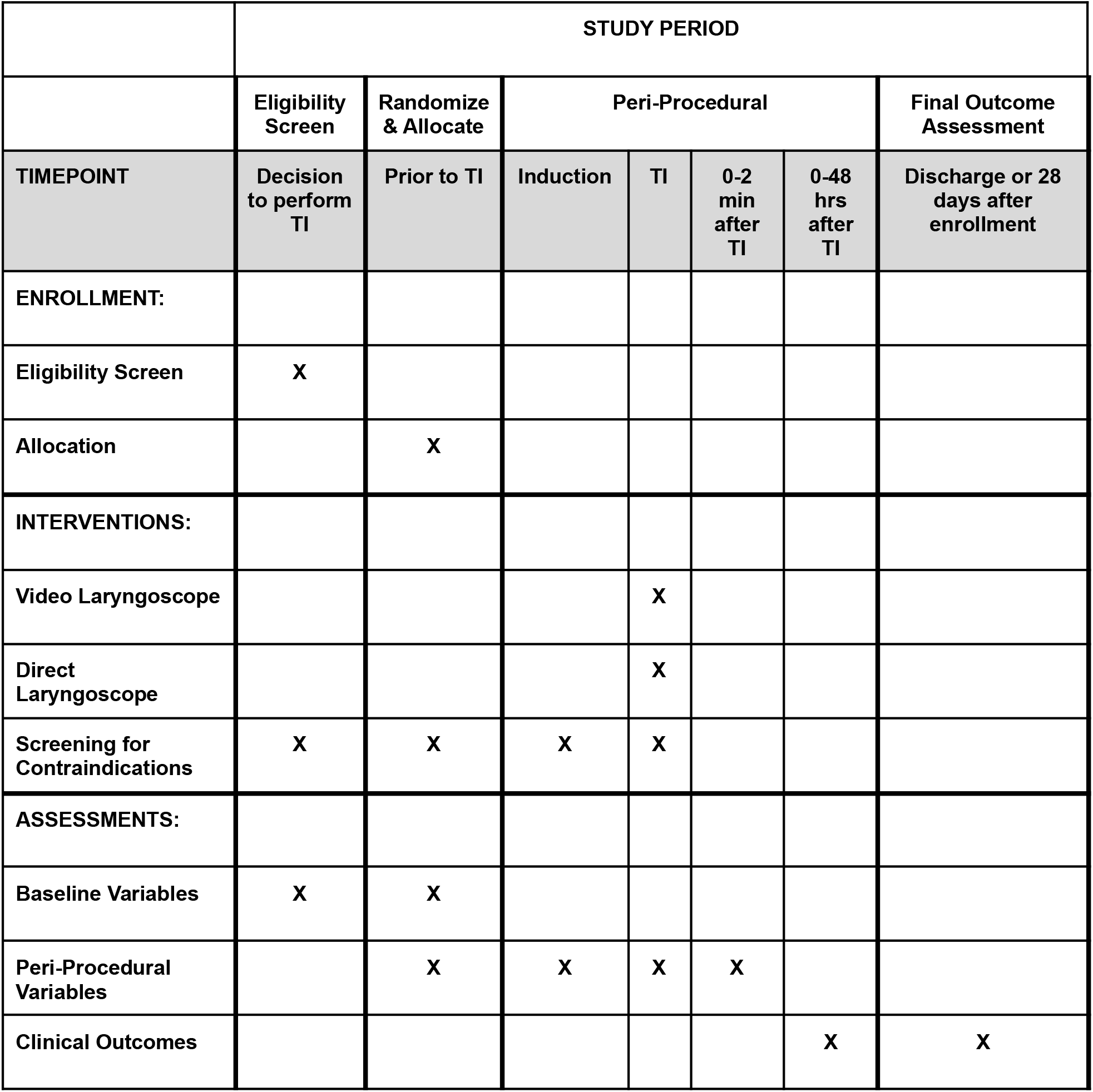
Schedule of enrollment, interventions, and assessments in the DEVICE trial. TI, tracheal intubation.

### Patient and Public Involvement

Materials used to communicate details of the study with patients and family members were developed with input from the Vanderbilt Community Advisory Council. Study authors will disseminate the results of this study online and via social media in forms suitable for public understanding.

### Study Design

The <u>D</u>ir<u>E</u>ct Versus <u>VI</u>deo Laryngos<u>C</u>op<u>E</u> (DEVICE) trial is a pragmatic, multicenter, unblinded, parallel-group, randomized trial comparing use of a video laryngoscope to use of a direct laryngoscope for the first attempt at emergency tracheal intubation among critically ill adults in the ED and ICU. The primary outcome is successful intubation on the first attempt. An independent data and safety monitoring board (DSMB) is monitoring the progress and safety of the trial. Study institutions and investigators are listed in the supplementary file, section 2.

### Study Population

The inclusion criteria for this study are:

1. Patient is located in a participating unit
2. Planned procedure is orotracheal intubation using a laryngoscope.
3. Planned operator is a clinician expected to routinely perform tracheal intubation in the participating unit.

The exclusion criteria for the study are:

1. Patient is known to be less than 18 years old
2. Patient is known to be pregnant.
3. Patient is known to be a prisoner.
4. Immediate need for tracheal intubation precludes safe performance of study procedures.
5. Operator has determined that use of a video laryngoscope or use of a direct laryngoscope is required or contraindicated for the optimal care of the patient.

### Randomization and Treatment Allocation

Patients are randomized in a 1:1 ratio to undergo intubation using a video laryngoscope or using a direct laryngoscope for the first attempt in permuted blocks of variable size, stratified by study site. Study-group assignments are generated using a computerized randomization sequence, placed in sequentially numbered opaque envelopes, and distributed to enrolling sites. Before opening the envelope, the operator determines that the patient meets eligibility criteria, records the predicted difficulty of intubation (“easy”, “moderate”, or “difficult”) and selects the blade shape the operator plans to use if the patient is randomized to the video laryngoscope group (“hyperangulated” or “non-hyperangulated / standard geometry”). The operator or delegate then opens the envelope. Patients are enrolled once the envelope is opened to reveal the study group assignment. After enrollment and randomization, patients, treating clinicians, and study personnel are not blinded to study group assignment.

### Study Interventions

#### Video Laryngoscope Group

For patients assigned to the video laryngoscope group, operators are instructed to use a video laryngoscope on the first laryngoscopy attempt. A video laryngoscope is defined as a laryngoscope with a camera and a video screen. Trial protocol does not dictate the brand of video laryngoscope or the geometry of the laryngoscope blade (e.g. hyperangulated vs. non-hyperangulated), but these details will be recorded. Operators are encouraged, but not required, to view the video screen during laryngoscopy (“indirect laryngoscopy”) and tracheal intubation.

#### Direct Laryngoscope Group

For patients assigned to the direct laryngoscope group, operators are instructed to use a direct laryngoscope on the first laryngoscopy attempt. A direct laryngoscope is defined as a laryngoscope without a camera and a video screen. Trial protocol does not dictate the brand of direct laryngoscope or the geometry of the laryngoscope blade (e.g. curved [Macintosh] vs. straight [Miller]), but these details will be recorded.

#### Co-Interventions and Subsequent Attempts at Laryngoscopy and Intubation

Study group assignment determines only the type of laryngoscope (video vs direct) used on the first laryngoscopy attempt. If determined to be required to ensure optimal care of the patient, treating clinicians may use any device at any time, regardless of study group assignment. Cases in which clinicians use a laryngoscope discordant with randomized assignment on the first intubation attempt will be documented and tracked. All aspects of the intubation procedure, except the type of laryngoscope used on the first attempt, are at the discretion of treating clinicians, including selection of sedative and neuromuscular blocking medications, patient positioning, approach to pre-oxygenation, use of a bougie or a stylet, and endotracheal tube size. Best practices in tracheal intubation will be encouraged according to clinical protocols at the study sites. The trial intervention ends after the first attempt at laryngoscopy. If the first attempt is unsuccessful, the operator may use any method of intubation on subsequent intubation attempts, including use of a direct laryngoscope in the video laryngoscope group or use of a video laryngoscope in the direct laryngoscope group. The type of laryngoscope used during the initial and final laryngoscopy attempt will be collected and reported.

#### Data Collection

A trained observer, not directly involved with the intubation procedure, collects data for key peri-procedural outcomes. These outcomes include successful intubation on the first attempt, time interval between laryngoscopy and successful intubation, the oxygen saturation and systolic blood pressure at induction, the lowest oxygen saturation and systolic blood pressure between induction and 2 minutes after successful intubation, and new or increased vasopressor administration between induction and 2 minutes after successful intubation. Observers may be clinical personnel on the enrolling unit (e.g., physician, nurse, or pharmacist) or research study personnel.

Immediately following the intubation procedure, the operator completes a paper data collection form to record the approach to preoxygenation, oxygenation and ventilation between induction and laryngoscopy, the brand of laryngoscope used, the blade shape, the Cormack-Lehane grade of laryngeal view^35^, use of the video screen to visualize the larynx (if applicable), use of a bougie or a stylet, reasons for failure to intubate on the first attempt (if applicable), intubation approaches on subsequent attempts, difficult airway characteristics observed before or during the procedure (facial trauma, small mouth opening, limited neck mobility, cervical collar, large neck, obesity, fluids obscuring view of vocal cords, upper airway obstruction or edema), and complications of intubation (witnessed pulmonary aspiration, esophageal intubation, injury to airways, injury to teeth, cardiac arrest between induction and 2 minutes following intubation). Operators record their specialty, training level, and estimates of the number of previous intubations they have performed and the number of previous intubations they have performed using a direct laryngoscope.

Study personnel at each site review the medical record to collect data on baseline patient characteristics, pre- and post-laryngoscopy management, and clinical outcomes at 28 days after enrollment.

The following variables are collected:

1. <u>Baseline</u>: Age, sex, height, weight, race, ethnicity, Acute Physiology and Chronic Health Evaluation II (APACHE II) score^36^, active medical problems at the time of enrollment, comorbidities, indication for intubation, vasopressor receipt in the hour prior to enrollment, highest FIO_2_ in the hour prior to enrollment, lowest SpO_2_/FIO_2_ (or PaO_2_/FIO_2_) ratio in the hour prior to enrollment, pre-procedural Glasgow Coma Scale (GCS) score^37^, oxygen delivery device at enrollment, assessment of the likelihood of a difficult intubation, presence of difficult airway characteristics (limited mouth opening, small mandible, large tongue, short neck, large neck circumference, limited anatomic neck mobility, cervical immobilization due to trauma, obesity), operator’s level of training and specialty, operator’s prior intubation experience.
2. <u>Peri-procedural</u>: Lowest SpO_2_ from enrollment to induction, approach to and duration of pre-oxygenation, time of sedative administration, sedative agent and dose administered, neuromuscular blocking agent and dose administered, SpO_2_ and systolic blood pressure at the time of induction, approach to oxygen administration and ventilation between induction and the first attempt at laryngoscopy, time of start of first laryngoscopy attempt, laryngoscope used on first attempt (model, blade size, blade shape), use of video screen (if applicable) on the first laryngoscopy attempt, best Cormack-Lehane grade of view^35^ on the first laryngoscopy attempt, presence of body fluid obstructing view of the larynx, presence of upper airway obstruction or edema, number of intubation attempts (number of times the laryngoscope entered the mouth, number of times the bougie entered mouth [if applicable], number of times the endotracheal tube entered the mouth), reason for failure of the first intubation attempt (if applicable), procedural adjustments made for the final intubation attempt, esophageal intubation, injury to teeth, operator-reported pulmonary aspiration between induction and intubation, time of successful tracheal intubation, endotracheal tube size, lowest SpO_2_ from induction until 2 minutes after intubation, lowest systolic blood pressure from induction until 2 minutes after intubation, new or increased vasopressor administration from induction until 2 minutes after intubation, cardiac arrest from induction until 2 minutes after intubation not resulting in death within 1 hour of induction, cardiac arrest from induction until 2 minutes after intubation resulting in death within 1 hour of induction.
3. <u>24 hours after enrollment</u>: new pneumothorax detected in the first 24 hours after induction, vasopressor receipt at 24 hours after induction, SpO_2_ at 24 hours after induction, FIO_2_ at 24 hours after induction, positive end-expiratory pressure (PEEP) at 24 hours after induction, systolic blood pressure at 24 hours after induction.
4. <u>In-Hospital Outcomes</u>: Ventilator-free days in the first 28 days, ICU-free days in the first 28 days, and in-hospital mortality at 28 days. Definitions for ICU-free days and ventilator-free days are provided in the supplementary file, sections 3 and 4.

#### Primary Outcome

The primary outcome is successful intubation on the first attempt. Successful intubation on the first attempt is defined as placement of an endotracheal tube in the trachea following a single insertion of a laryngoscope blade into the mouth and *either* a single insertion of an endotracheal tube into the mouth *or* a single insertion of a bougie into the mouth followed by a single insertion of an endotracheal tube into the mouth.

Data for the assessment of the primary outcome are collected by a trained independent observer using a structured data collection form that records the number of insertions of the laryngoscope blade, bougie (if used), and endotracheal tube into the patient’s mouth. In the event that data from the independent observer are missing, data from the operator’s self-report of successful intubation on the first attempt will be used.

#### Secondary Outcome

The secondary outcome is the incidence of severe complications occurring between induction and 2 minutes following successful intubation. Severe complications are defined as one or more of the following:

- Severe hypoxemia (lowest oxygen saturation measured by pulse oximetry < 80%);
- Severe hypotension (systolic blood pressure < 65 mm Hg or new or increased vasopressor administration);
- Cardiac arrest not resulting in death
- Cardiac arrest resulting in death

Cardiac arrest will be considered to have resulted in death if a patient who experienced cardiac arrest between induction and 2 minutes after intubation died within the 1 hour following intubation.

### Exploratory Outcomes

#### Exploratory procedural outcomes are as follows

- Duration of laryngoscopy and tracheal intubation. This is defined as the interval (in seconds) between the first insertion of a laryngoscope blade into the mouth and the final placement of an endotracheal tube or tracheostomy tube in the trachea.
- Number of laryngoscopy attempts
- Number of attempts to cannulate the trachea with a bougie or endotracheal tube
- Successful intubation on the first attempt without a severe complication
- Reason for failure to intubate the trachea on the first attempt, which include:
  - Inadequate view of the larynx
  - Inability to intubate the trachea with an endotracheal tube
  - Inability to cannulate the trachea with a bougie
  - Attempt aborted due to a change in patient condition (e.g. worsened hypoxemia, hypotension, bradycardia, vomiting, bleeding)
  - Technical failure of the laryngoscope (e.g. battery, light source, camera, screen)
  - Other
- Operator-reported aspiration

Exploratory safety outcomes are as follows:

- Esophageal intubation
- Injury to the teeth

Exploratory clinical outcomes are as follows:

- ICU-free days in the first 28 days
- Ventilator-free days in the first 28 days
- 28-day all-cause in-hospital mortality

### Sample Size Estimation

The minimum clinically important difference in successful intubation on the first attempt that would be needed to justify routine use of a video laryngoscope rather than a direct laryngoscope in the ED and ICU is uncertain. The current trial is designed to detect a 5% absolute difference between groups in the incidence of successful intubation on the first attempt. An absolute difference of 5% in successful intubation on the first attempt is similar to or smaller than the difference used in the design of prior airway management trials and is considered by airway management experts to be clinically meaningful.^21,28,38,39^ Assuming (1) an incidence of successful intubation on the first attempt of 80% in the direct laryngoscope group, (2) 90% statistical power, (3) a two-sided alpha of 0.05, and (4) enrollment at 16 sites with an intra-cluster correlation for the primary outcome of 0.05, we calculated that detecting a 5% absolute increase in the incidence of successful intubation on the first attempt would require enrollment of 1,920 patients (960 per group). Anticipating missing data for up to 4% of enrolled patients, we will plan to enroll a total of 2,000 patients (1,000 per group).

### Data and Safety Monitoring Board (DSMB) and Interim Analysis

A DSMB composed of experts with backgrounds in emergency medicine, pulmonary and critical care medicine, anesthesiology, bioethics, and biostatistics has overseen the design of the trial and is monitoring its conduct. The DSMB will review a single interim analysis prepared by the study biostatistician at the anticipated halfway point of the trial, after enrollment of 1,000 patients. The stopping boundary for efficacy was pre-specified as a P-value of 0.001 or less, using a chi-square test, for the difference in the incidence of the primary outcome between groups. This conservative Haybittle–Peto boundary was selected to allow the final analysis to be performed using an unchanged level of significance (P < 0.05). The DSMB retains the authority to stop the trial at any point, request additional data or interim analyses, or request modifications of the study protocol to protect patient safety. Trial safety monitoring and handling of adverse events are described in detail in the supplementary file, section 5. Patient privacy and data storage details are listed in the supplementary file, section 6.

### Statistical Analysis Principles

Analyses will be conducted following reproducible research principles using R (R Foundation for Statistical Computing, Vienna, Austria).^40^ We will present summary tabulations by treatment group. For categorical variables, the number and proportion of patients will be presented. For continuous variables, the mean and standard deviation or median and interquartile range will be presented, as appropriate.

We will analyze a single pre-specified primary outcome and a single pre-specified secondary outcome using a chi-square test. Consistent with recommendations of the Food and Drug Administration^41^ and the European Medicines Agency^42^, each will be tested using a two-sided P value with a significance level of 0.05. The primary analysis will occur in an intent-to-treat fashion among all patients randomized, excluding only those patients whose data was withdrawn from the study. For all other analyses except safety analyses, emphasis will be placed on the estimate of effect size with 95% confidence intervals, as recommended by the *International Committee of Medical Journal Editors*^*43*^, and no corrections for multiple comparisons will be performed.

### Main Analysis of the Primary Outcome

The main analysis will be an unadjusted, intention-to-treat comparison of successful intubation on the first attempt between patients randomized to the video laryngoscope group and patients randomized to the direct laryngoscope group, using a chi-square test. The difference in proportions, the associated 95% confidence interval, and a p value for the primary outcome will be presented.

### Secondary Analyses of the Primary Outcome

#### Multivariable modeling to account for covariates

To account for relevant covariates, we will develop a generalized linear mixed effects model using a logit link function with the primary outcome as the dependent variable, study site as a random effect, and fixed effects of study group and the following pre-specified baseline covariates: age, sex, body-mass index, operator experience quantified as the operator’s total number of prior intubations, and location of intubation (ED vs ICU). All continuous variables will be modeled assuming a nonlinear relationship to the outcome using restricted cubic splines with between 3 and 5 knots.

#### Effect Modification

We will examine whether pre-specified baseline variables modify the effect of study group assignment (video laryngoscope vs direct laryngoscope) on the primary outcome using a formal test of statistical interaction in a generalized linear mixed effects model with the primary outcome as the dependent variable, study site as a random effect, and fixed effects of study group, the pre-specified proposed effect modifier, and the interaction between the two. For categorical variables, we will present the odds ratio and 95% confidence intervals within each pre-specified subgroup. Continuous variables will not be dichotomized for analysis of effect modification but may be dichotomized for data presentation. In accordance with the Instrument for assessing the Credibility of Effect Modification Analyses (ICEMAN) recommendations^44^, we have prespecified the following limited number of baseline variables as potential effect modifiers and the hypothesized direction of effect modification for each:

1. <u>Patient location (ED vs ICU)</u>. We hypothesize that patient location will not modify the effect of study group assignment on the primary outcome.
2. <u>Traumatic injury (Yes vs No)</u>. We hypothesize that traumatic injury will modify the effect of study group assignment on the primary outcome, with a greater increase in the incidence of successful intubation on the first attempt with use of a video laryngoscope compared with a direct laryngoscope among patients with traumatic injury compared to patients without traumatic injury.
3. Body mass index (kg/m^2^). We hypothesize that body mass index will modify the effect of study group assignment on the primary outcome, with a greater increase in the incidence of successful intubation on the first attempt with use of a video laryngoscope compared with a direct laryngoscope among patients with higher body mass index as compared to patients with lower body mass index. This hypothesis of effect modification is supported by a non-significant trend toward effect modification in a meta-analysis of multiple prior randomized trials.^9^
4. <u>Operator’s pre-enrollment assessment of the anticipated difficulty of intubation (Easy; Moderate; Difficult; Not Recorded)</u>. We hypothesize that the operator’s pre-enrollment assessment will modify the effect of study group assignment on the primary outcome, with a greater increase in the incidence of successful intubation on the first attempt with use of a video laryngoscope compared with a direct laryngoscope among patients assessed as Difficult or Moderate compared to Easy. This hypothesis of effect modification is supported by significant effect modification in a meta-analysis of multiple prior randomized trials.^9^
5. Operator experience at the time of enrollment.
  1. <u>Total number of previous intubations performed by operator</u>. We hypothesize that the total number of previous intubations performed by the operator will modify the effect of study group assignment on the primary outcome, with a greater increase in the incidence of successful intubation on the first attempt with use of a video laryngoscope compared with a direct laryngoscope among operators with fewer previous intubations compared to operators with a greater number of previous intubations. This hypothesis of effect modification is supported by significant effect modification observed in a prior randomized trial among critically ill adults, but differs from a meta-analysis including trials of intubation in the operating room that did not observe effect modification based on the operator’s prior experience.^9,28^
  2. <u>Proportion of previous intubations performed by the operator using a direct laryngoscope</u>. We hypothesize that the proportion of previous intubations performed by the operator using a direct laryngoscope will modify the effect of study group assignment on the primary outcome, with a greater increase in the incidence of successful intubation on the first attempt with use of a video laryngoscope compared with a direct laryngoscope among operators with a lower proportion of previous intubations performed by the operator using a direct laryngoscope compared to operators with a higher proportion of previous intubations performed by the operator using a direct laryngoscope.

We will also perform an effect modification analysis for the primary outcome that includes a three-way interaction between study group, total number of previous intubations performed by the operator, and proportion of previous intubations performed by the operator using a direct laryngoscope.

### Sensitivity Analyses of the Primary Outcome

We will assess the robustness of the findings of the primary analysis in a number of sensitivity analyses. First, because operators may choose to deviate from the assigned laryngoscope for the safety of the patient, we will repeat the primary analysis, but will consider patients for whom the operator crossed over on the first attempt from the assigned laryngoscope type to the non-assigned laryngoscope type not to have experienced successful intubation on the first attempt. Second, we will repeat the primary analysis among only patients for whom data on the primary outcome from the independent observer is available (i.e., excluding cases in which operator self-report was the sole source of information for the primary outcome). Third, because the operator’s prior experience with each type of laryngoscope may affect the likelihood of success with a video laryngoscope compared with a direct laryngoscope, we will repeat the primary analysis among only cases in which the proportion of prior intubations the operator has performed using a direct laryngoscope is between 0.25 and 0.75.

### Analysis of the Secondary Outcome

For the secondary outcome, severe complications occurring between induction and 2 minutes following intubation, we will perform an unadjusted, intention-to-treat comparison of patients randomized to the video laryngoscope group versus patients randomized to the direct laryngoscope group, using a chi-square test.

### Analyses of Exploratory Outcomes

For all pre-specified exploratory outcomes, we will conduct unadjusted, intention-to-treat analyses comparing patients randomized to the video laryngoscope group versus patients randomized to the direct laryngoscope group. We will calculate absolute risk differences or differences in medians between groups with the associated 95% confidence intervals.

### Handling of Missing Data

We anticipate that no data on the primary outcome will be missing. When data are missing for the secondary or exploratory outcomes, we will perform complete-case analysis, excluding cases where the data for the analyzed outcome are missing. There will be no imputation of missing data for these outcomes. In adjusted analyses, missing data for covariates will be imputed using multiple imputations.

### Trial status

The <u>D</u>ir<u>E</u>ct Versus <u>VI</u>deo Laryngos<u>C</u>op<u>E</u> (DEVICE) trial is a prospective, multi-center, non-blinded randomized clinical trial comparing use of a video laryngoscope to use of a direct laryngoscope for the first attempt at tracheal intubation of critically ill adults in the ED and ICU. Patient enrollment began on 16 March 2022 and is being conducted in 6 EDs and 10 ICUs in the United States.

## Ethics and Dissemination

### Waiver of Informed Consent

Critically ill patients undergoing tracheal intubation in the ED or ICU are at significant risk for morbidity and mortality from their underlying illness. Most patients undergoing tracheal intubation in routine clinical care are intubated using either a video laryngoscope or a direct laryngoscope on the first attempt. Any benefits or risks of these two approaches are experienced by patients undergoing tracheal intubation in clinical care, outside the context of research. As a requirement for enrollment in the DEVICE trial, the patient’s treating clinician must believe that either a video laryngoscope or a direct laryngoscope would be a safe and reasonable approach for the patient (otherwise the patient is excluded). Therefore, making the decision between the two approaches randomly (by study group assignment) rather than by a clinician who thinks either approach is safe and reasonable for the patient is expected to pose no more than minimal additional risk.

Obtaining informed consent for participation in the study would be impracticable. The majority of patients undergoing emergency tracheal intubation lack decisional capacity due to their underlying critical illness and surrogate decision makers are frequently absent. Further, emergency tracheal intubation is a time-sensitive procedure with only minutes between the decision to perform intubation and the completion of the procedure. Meaningful informed consent could not be executed in this brief window and attempting to obtain informed consent would lead to potentially deleterious and unethical delays in intubation which would increase the risk of hypoxemia, hypotension, and periprocedural cardiac arrest.

Because the study involves minimal incremental risk, the study would not adversely affect the welfare or privacy rights of the participant, and obtaining informed consent would be impracticable, a waiver of informed consent was requested from and approved by the single institutional review board at Vanderbilt University Medical Center (reference number 211272). This is consistent with previous randomized trials comparing alternative approaches to tracheal intubation commonly used in clinical care.^28,38,39,45–50^ This approach was approved by the US Department of Defense (DoD) Defense Health Agency Human Research Protection Office (EIRB# 944893).

### Information for Patients and Families

Information regarding the study is made available to patients and families using a patient and family information sheet. The patient and family information sheet contains information on the purpose of the trial, study procedures, risks and discomforts, benefits, use of protected health information, confidentiality, and investigator contact information. The Defense Health Agency Human Research Protection Office determined that this procedure meets the requirements of 32 CFR 219 and DODI 3216.02_AFI40-402. At centers with a significant population of non-English speaking patients, the patient and family information sheet has been translated into Spanish and Somali languages and is made available to those patients.

### Protocol Changes

Any further amendments to the protocol will be recorded on ClinicalTrials.gov as per SPIRIT guidelines. See the supplementary file, section 7, for details on how protocol changes will be handled.

### Dissemination Plan

Trial results will be submitted to a peer-reviewed journal and will be presented at one or more scientific conferences.

## Data Availability

All data that will be produced in the present study are available upon reasonable request to the authors

## Authors’ Contributions

Approved the final version of this manuscript and critical revision of the manuscript for important intellectual content: all authors. Study concept and design: MEP, BED, SAT, SGS, BJL, DRG, WHS, TWR, AAG, JDC, MWS. Acquisition of data: all authors. Drafting of the manuscript and study supervision: MEP, BED, SAT, JDC, MWS.

